# Using machine learning to identify subgroups with the highest expected benefit in a population-based water, sanitation, handwashing, and nutrition intervention

**DOI:** 10.1101/2025.06.17.25329796

**Authors:** Caitlin Hemlock, Laura H. Kwong, Lia C.H. Fernald, Alan E. Hubbard, John M. Colford, Fahmida Tofail, Md. Mahbubur Rahman, Sarker Parvez, Stephen P. Luby, Andrew N. Mertens

**Affiliations:** Department of Environmental and Occupational Health, University of Washington, Seattle, WA, USA; Division of Epidemiology, School of Public Health, University of California, Berkeley, Berkeley, CA, USA; Division of Environmental Health Sciences, School of Public Health, University of California, Berkeley, Berkeley, CA, USA; Division of Community Health Sciences, School of Public Health, University of California, Berkeley, Berkeley, CA, USA; Division of Biostatistics, School of Public Health, University of California, Berkeley, Berkeley, CA, USA; Nutrition Research Division, icddr,b, Dhaka-1212, Bangladesh; Environmental Health and WASH, Health System and Population Studies Division, icddr,b, Dhaka-1212, Bangladesh; Global Health and Migration Unit, Department of Women’s and Children’s Health, Uppsala University, Sweden; Children’s Health and Environment Program, Child Health Research Centre, The University of Queensland, South Brisbane, QLD, Australia; Division of Infectious Diseases and Geographic Medicine, Stanford University, Stanford, CA USA

## Abstract

**Background:** Understanding who benefits most from investments in water, sanitation, and hygiene (WaSH) interventions can elucidate causal pathways, uncover complex interactions between population characteristics and interventions, and inform targeted implementation. We applied machine learning to identify and describe households of children that benefited most from WaSH and nutrition interventions.

**Methods:** We used causal forests and baseline characteristics of pregnant women enroled in a trial in Bangladesh (2013-2015) to test for heterogenous treatment effects of the primary trial outcomes at two years (length-for-age Z-score [LAZ-score] and diarrhoea prevalence) and one secondary outcome (child development [EASQ Z-score]) for each treatment-outcome combination. We split households into three groups based on predicted treatment effect magnitude and compared characteristics of those that benefitted the most (Tercile 3) versus the least (Tercile 1).

**Results:** Heterogeneity was detected in the effect of Sanitation on EASQ Z-score, compared to Control; children in Tercile 3 were estimated to gain 0.51 SD (95% CI: 0.35, 0.67) whereas children in Tercile 1 were estimated to have no benefit. At baseline, households of children in Tercile 3 were more likely to report that chickens always entered the house (85% vs. 4%) and had animal feces observed in the child’s play area (84% vs. 18%) when compared with Tercile 1. Tercile 3 households also owned less land and assets and lived further from Dhaka, any population center, or a market. We did not detect heterogeneity for any other treatment-outcome comparison.

**Conclusions:** We did not detect heterogeneity in any treatment arms for the outcomes of diarrhoea or LAZ-score, showing that children from all backgrounds benefit from effective interventions equally based on household characteristics. We found heterogeneity in the effect of receiving sanitation improvements on child development, where poorer households located in more remote areas and potentially with higher levels of animal fecal contamination had the highest expected benefit.

## BACKGROUND

Many interventions that have produced outcome effectiveness on a small scale, including water, sanitation, and hygiene-related interventions (1), have not demonstrated the same effects when scaled to entire populations. Factors potentially influencing this include inequities in implementation and uptake and underlying heterogeneity within the targeted population (2).

However, recent statistical advances have provided a platform to uncover potential heterogeneous effects of interventions and may supply insight into the complex dynamics of where, when, and among whom such interventions have maximal effects. Identifying household or individual characteristics that result in heterogeneous effects of an intervention may be beneficial for implementing population-scale interventions (3), as well as generating an understanding of the differences in results between settings and further influencing policy by identifying the most cost-effective groups to target.

Traditional approaches to detect heterogeneous treatment effects involve pre-specifying one or more variables for subgroup analyses. Posthoc subgroup analyses are frowned upon due to the potential for bias by analytic investigators and for type I (false positive) and type II (false negative) errors (4). However, drawbacks to this prespecified approach include an *a priori* understanding of the causal mechanisms interacting with treatment that may or may not be accurate or comprehensive and not allow for novel discoveries. An alternative approach is causal forests, which leverages recursive partitioning with random forests to estimate the conditional average treatment effect (CATE), or the average treatment effect conditional on sets of variables (5,6). These methods test if a set of prespecified, observed variables resulted in heterogeneous effects; this prevents potential data mining while still uncovering complex interactions with the treatment (7). The use of causal forests and other machine learning algorithms to detect treatment effect heterogeneity can be a powerful way to detect subgroups defined by different combinations of covariates and pinpoint populations that may benefit most from interventions.

These methods have been used more commonly in fields such as economics, pharmacoepidemiology, and health policy; for example, Chernozhukov *et al.* found that out of a set of prespecified effect modifiers, distributing incentives among villages with the lowest fraction of immunized children had the greatest impact on immunization takeup (8) and Athey *et al.* determined that unmarried adolescents were more receptive to discounts on long-acting reversible contraceptives (9). As the field of implementation science grows and its principles are applied to global health problems (10), including inadequate WASH (1), applications of these methods may help both discover important modifiers of intervention impact and identify potential targeting strategies to maximize effectiveness in resource-constrained settings.

The WASH Benefits trials conducted in Bangladesh and Kenya tested the effects of water, sanitation, and handwashing (WASH) interventions, previously shown to be efficacious in reducing illness (11), in a population-based setting. The goal was to reduce illness and subsequently improve child growth and development (12,13). The interventions created an enabling environment by delivering water, sanitation, and hygiene (WASH) hardware and behaviour-change messaging, in combination with nutrition supplementation in a subset of study population, with the targeted outcome of reducing diarrhoeal infections in young children and improving their growth and development (14). While the goal of the trial was to assess independent and combined effects of WASH and nutrition interventions on the reduction of diarrhoea and linear growth faltering, investigators also aimed to develop low-cost interventions that could be scaled if the trial successfully achieved the primary outcomes. In Bangladesh, the water-only, sanitation-only, and hygiene-only interventions reduced diarrhoea by 32-39% (around two percentage points). However, there was no synergism in the combined water, sanitation, and handwashing intervention arm compared to the individual intervention arms, and no WASH intervention arm improved growth without the nutritional intervention present. The study also found strong effects of all interventions across a range of child development measures, but with no synergism in any combined treatment arms (15). With null results for diarrhoea in Kenya and growth in both trials for the WASH arms, many in the WASH field have called on “transformative WASH” in future research, which includes higher-cost, community-level interventions (16). Given that the household-level interventions in the trials were rigorously piloted, we posit that there may have been subgroups of households that were still substantially impacted by the interventions (e.g. households with worse sanitation infrastructure at baseline or households with more resources and better able to implement behaviour change), while the majority were not, resulting in the overall null effect for growth. Understanding subgroups of households that were impacted more by the interventions may also lend insight into the mechanism by which improvements were mediated.

Although we may not see substantial heterogeneity in the treatment effects such as those seen in pharmaceutical interventions (where interventions are dynamic and may be harmful in some groups), given that the interventions were unlikely to produce negative effects and the average treatment effects for diarrhoea and growth were small, some evidence points to the potential for heterogeneous effects. A recent article found that the average treatment effect on diarrhoea in the combined WASH arms in this trial was highest among the poorest quartile of the study population specifically during the rainy season (17), while another article found that any WASH intervention reduced diarrhoea more during periods of high precipitation or temperature (18).

One reason could be greater adoption of the hardware and messages delivered by community health promoters was higher in certain subgroups, which was already found in household socioeconomic status differences in uptake; the poorest households demonstrated the greatest change in uptake from baseline (19). It also may have been driven by heterogeneity in WASH coverage in the target population prior to the intervention (12,13); around half of the households in the population already owned a water storage device or a latrine (one-third of which had a functional water seal) which was hardware delivered by the study to improve WASH conditions. There also may have been other pathways by which primary outcome gains were achieved in certain sub-populations (e.g. improvement in maternal factors in worse-off mothers directly affecting child outcomes outside of WASH-related pathways (20)) or negated (incomplete coverage of fecal contamination pathways in households with high levels of contamination (21)).

Our overall objective was to understand which, if any, observed baseline characteristics generated heterogeneous treatment effects for diarrhoeal prevalence, child growth, and child development at endline among enroled households across each of the treatment arms in the WASH Benefits Bangladesh trial compared to the control arm. Our specific objectives included identifying individual-, household-, or village-level characteristics that caused the largest treatment effects using causal forests, which may be informative for understanding causal pathways and implementing future household-level water, sanitation, and hygiene interventions in similar contexts.

## METHODS

### Original intervention trial

#### Study design

The WASH Benefits Bangladesh study was a two-year, cluster randomized controlled trial conducted in the Gazipur, Kishoreganj, Mymensingh, and Tangail districts of Bangladesh from 2012-2015 (ClinicalTrials.gov registration number: NCT01590095). The trial was designed to measure the impact of individual and combined nutrition, water, sanitation, and handwashing interventions on health outcomes in young children. Details of the interventions have been previously published (22), as have papers reporting the main effects of the interventions (12,15). Briefly, the six intervention arms consisted of an insulated storage container for drinking water and chlorine tabs for water treatment (Water [W]); provision of a double-pit latrine with a functional water seal, a child potty if there was already a child younger than 3 years old, and a tool for the removal of feces that was designed and piloted in this context (23) (Sanitation [S]); two handwashing stations, storage bottle for soapy water, and laundry detergent sachets for preparation of soapy water (Handwashing [H]); a combined intervention integrating all interventions from the W, S, and H arms (WSH); lipid nutrient supplementation (Nutriset, France) and promotion of breastfeeding and diverse complementary foods (Nutrition [N]); and a combined intervention integrating N + WSH arms. Compounds in the double-sized control arm did not receive any interventions.

#### Enrolment

Study staff recruited pregnant women (targeting women in their first or second trimester) in geographical clusters of eight compounds (groups of patrilineal households placed around a central courtyard, usually with a shared water source and latrine). In the intervention arms, community health promoters (CHPs) visited the compounds at least weekly during the first six months, and then biweekly for the remaining study period to deliver behaviour change messaging associated with each intervention. CHPs were hired for the purposes of the study and were required to have eight years of formal education, live within walking distance of enroled compounds, and pass a written and oral examination.

#### Randomization/sampling

Clusters were stratified by geographical location and block randomized. Further details of the recruitment and sampling methods, inclusion and exclusion criteria, and enrolment processes are detailed in the published results of the main trial (12,15).

#### Data collection

At baseline, enumerators conducted a census of each target child’s compound and conducted a comprehensive survey on the child’s household with information related to household demographics, water, sanitation, hygiene, and animal ownership and husbandry characteristics. Global positioning system (GPS) coordinates of the compound were also collected. At midline and endline, when the target children were approximately one and two years, enumerators visited compounds to collect data on adherence and outcomes.

### Outcomes of interest

We tested treatment effect heterogeneity for outcome variables of interest along the hypothesized causal chain for the interventions (Figure 1). The specific measures for this paper include the primary outcomes reported in the main trial, diarrhoea and linear growth, and one secondary outcome, child development, all measured during the endline assessment. We used a complete case analysis with all outcome variables.

**Figure 1.**
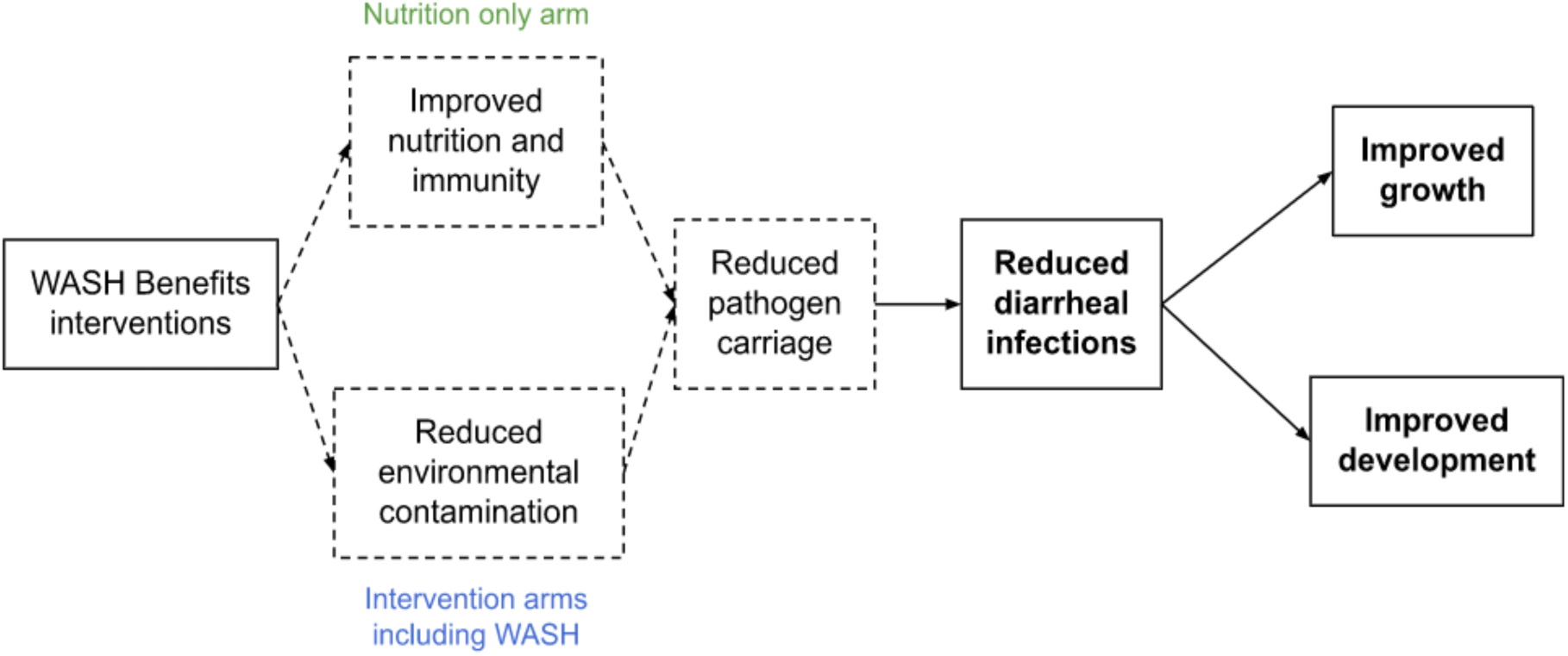
*A priori* causal pathway for WASH Benefits for achieving proximal and distal outcomes through interventions. Note: Heterogeneity only test among bolded outcomes

For diarrhoea, caregivers were asked to recall if any children in the compound <3 years had diarrhoea, defined using the WHO definition of 3 or more loose stools within 24 hours, in the past 7 days.

For linear growth, pairs of trained anthropometrists measured the index child’s recumbent length (accurate to 0.1 cm) in triplicate following standard protocols for anthropometric outcomes measurement.(24) The median of the three measurements was used to calculate length-for-age Z score (LAZ) using WHO 2006 growth standards (25). Child age was determined using birth dates, verified using vaccination cards when possible.

For child development, staff administered several assessments at endline to index children. For this analysis, we focused on the Extended Ages and Stages Questionnaire (EASQ), as it is a screening assessment that is widely utilized for assessing child development in low resource settings. The EASQ is primarily a parent-reported measure of child developmental progress and was adapted and validated for use in Bangladesh (adding direct assessment to 25% of the items). Trained enumerators asked parents questions about their child’s development and asked children to demonstrate certain behaviours (15). The adapted scale measured three out of five possible domains: child communication, gross motor, and personal-social skills, and combined the three domains into a combined measure, which was age-standardized to the control group distribution using 2-month age bands.

### Baseline variables of interest

We chose an initial set of variables collected in the baseline survey *a priori* and prespecified these as our predictors of interest (Supplementary Table 1). From this list, we dropped any variables with >10% missingness. All other missingness is handled in the estimation function in R using “missingness incorporated in attributes” (MIA) criterion (26), which creates indicator variables for missingness. We tested for strong correlations among remaining variables and observations using Pearson’s correlation coefficients (defining strong as *r* ≥ 0.7, moderate as *r* ≥ 0.3 and < 0.7, and weak as *r* < 0.3). All categorical variables collected in the survey were converted to dummy variables (binary indicator variables for each category). We also decided post hoc to drop outcomes of children with a high proportion of missingness across the selected baseline variables of interest (defined as >40% of variables missing).

### Parameter of interest

The target estimand for the original trial was the average treatment effect:

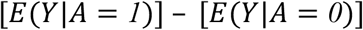

This is the difference between the average value of the outcome *Y* among those assigned to intervention *A* = *1* compared to those assigned to control *A* = *0*. In this analysis, we are interested in the average treatment effect *conditional* on baseline covariates, or the conditional average treatment effect. Our target estimand can be written as:

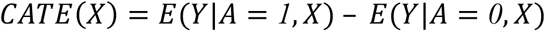

where E[*Y*|*A* = 1, *X*] is the conditional mean outcome observed if assigned to treatment (*A* = 1) given a vector of covariates *X* and E[*Y*|*A* = 0, *X*] is the conditional mean outcome observed if assigned to control given the same set of covariates. Under assumptions, the CATE(x) function identifies the counterfactual CATE, or *τ*(*x*) = *E*[*Y*(1) − *Y*(0) | *X* = *x*] or the difference in counterfactual means among group *X* = *x*. Given randomization and the empirical balance of baseline covariates across treatment arms previously published (12), we assumed that a nonparametric estimator of CATE(x) would be an unbiased estimate of *τ*(*x*)(27). Thus *τ*(*x*) is equivalent to the expected difference of the outcome if an individual was assigned to treatment versus control among the individuals where *X* = *x*.

### Estimation approach

We followed our prespecified analysis plan (https://osf.io/vr235/) to analyse heterogeneous treatment effects using the *grf* package in R. We estimated CATEs using causal forests, a random forest algorithm that creates splits where the difference of the CATE between leaves is maximized (6). In all analyses, we accounted for clustering using the trial cluster identification numbers and known treatment propensity (0.33 for each treatment arm-control arm comparison given the double-sized control arm in the trial).

#### Testing for heterogeneity

We constructed causal forests for each subsample of data created by combining each treatment arm with the control arm in turn, and subset to children with available outcome data. In the first stage, we constructed a forest using prespecified baseline variables of interest (after pruning for missingness and strong correlations as detailed above). Then, we refit the data in a second stage only on features that produced a reasonable number of splits in the forest (defined as variables with variable importance higher than the mean [see below]) (28). We tuned all parameters with cross-validation; this involved testing different values for different parameters, including the minimum number of observations in each tree leaf, the number of variables tried for each split, the fraction of data used to build each tree, maximum imbalance of the split, and the strength of the penalty applied when splits divide data into more uneven groups, and selecting the parameter values that minimized the Root Mean-Squared Error (RMSE) for the final causal forest. We used the variables selected for the second stage in all subsequent analyses.

Next, we tested the goodness-of-fit of each causal forest using the *test_calibration* function. The function approximates a proxy predictor of heterogeneity in the population (“best linear predictor”, as discussed by Chernozhukov *et al.*(8)) and outputs coefficients and p-values for two measures: the “differential forest prediction” and the “mean forest prediction”. The p-value for differential forest prediction acts as an omnibus test for the presence of heterogeneity; thus if the p-value is < 0.05, we can reject the null hypothesis that there is no heterogeneity. The p-value for mean forest prediction indicates whether the algorithm was able to predict an average treatment effect (H_0_: not able to predict); failing to reject (p ≥ 0.05) suggests that the causal forest may underestimate the true heterogeneity. We considered forests with p-values < 0.05 on the test for heterogeneity to be significant and concluded that heterogeneous treatment effects (HTE) exist.

#### Estimating group average treatment effects

After testing for significant HTE, we predicted individual treatment effects and estimated average treatment effects among subgroups of the population (called *Sorted Group Average Treatment Effects* [*GATES*]). We did this by first retraining each treatment-outcome causal forest using 10-fold cross-validation (keeping trial clusters together within folds) and predicting individual treatment effects on the out-of-bag samples, which maintains valid inference. After estimating individual treatment effects, we sorted individuals within each fold based on their predicted individual treatment effect (lowest to highest) and assigned each individual a tercile (reduced posthoc from quintiles to improve statistical power) where those with the smallest expected benefits were in the first tercile and those with the largest expected benefits were in the third tercile. We then calculated the treatment effect within each tercile (i.e., what we would have expected to see if we only treated that tercile of the population) using ordinary least squares regression. To estimate confidence intervals, we used a sandwich estimator to ensure heteroscedastic-consistent estimation of the covariance matrix, and bootstrapped 10,000 replicates of the predicted individual treatment effects from the trained forest within terciles. We corrected for multiple testing using the Romano-Wolf correction (29), which takes into account the dependence between tests. We considered tercile-specific treatment effects significantly different if the adjusted p-value < 0.05. We calculated GATES for causal forests of treatment-outcome combinations with heterogeneity p < 0.05, as well as those with p < 0.10 to illustrate predictions from causal forests with less predicted heterogeneity.

#### Important variables for predicting heterogeneity of treatment effects

To understand observed variables with the largest influence in creating treatment effect heterogeneity (i.e. the variables that influence *τ* the most), we performed a variable importance analysis. For random forest algorithms, variable importance is based on the frequency the variable is used across regression trees that make up the forest and the depth in the tree which they are used (variables at higher splits being more important) and is a measure of how influential the variable is in predicting the conditional average treatment effect. We extracted the variables from second-stage causal forests with significant heterogeneity and presented them in order of importance. We conducted a classification analysis (CLAN) on these variables among the outcomes with significant differences in expected benefits between terciles. We describe the differences in means and proportions in baseline characteristics among those with the largest (third tercile) and smallest (first tercile) benefit and tested for significant differences comparing the first and third terciles among the set of variables identified as important using univariate OLS regression and adjusted p-values using the Bonferroni correction (30).

We also estimated a linear combination of the top five important variables for selected causal forests using the *best_linear_projection* function to project *τ* on the variables. We scaled continuous predictor variables by the interquartile range so the coefficient output compared *τ* for those in the 75th percentile compared to those in the 25th percentile for the given variable; binary predictors compared *τ* for presence versus absence of the variable.

All analyses were performed in R Studio v4.2.1. Code is available at [Add github repo before publication].

## RESULTS

### Dataset

Among the 5551 pregnant women recruited at baseline, we removed 23 (0.4%) because of high missingness across chosen predictor variables. We matched baseline survey data on households to Year 2 outcome data among 8666 all children < 3 years with available diarrhoea prevalence, 4564 target children with available LAZ, and 4204 target children with available EASQ Z-scores. Of the 53 variables chosen from the baseline survey *a priori* (Supplementary Table 1), maternal height and population density variables were missing in more than 10% of observations and were dropped from the variable set. Baseline characteristics were balanced across treatment arms (Table S1). After encoding categorical variables to binary predictors for each level, we ended with 78 total predictors to use in our causal forests to predict treatment effect heterogeneity. Most variables were only weakly correlated with each other (Supplmentary Figure 1) with only 2.5% of pairwise comparisons having moderate correlation (*r* ≥ 0.3) and 0.3% of pairwise comparisons having strong correlation (*r* ≥ 0.7).

### Testing for treatment effect heterogeneity

The majority of causal forests constructed were able to accurately predict the outcomes (mean forest prediction p < 0.05), but most intervention-outcomes combinations did not exhibit heterogeneous treatment effects (differential forest prediction p ≥ 0.05) (Table 1). Heterogeneity was only detected for the EASQ Z-score outcome comparing the Sanitation arm to Control (p = 0.0028). The causal forests with the next strongest heterogeneity were those predicting the effects of the Handwashing intervention on EASQ Z-scores (p = 0.056) and the effects of the Water intervention on LAZ (p = 0.071).

**Table 1.** Testing causal forests for goodness of fit and heterogeneity.

|  | Main Trial Average<br>Treatment Effect<sup>1</sup> | Mean Forest<br>Prediction<sup>2</sup> | Differential Forest<br>Prediction<sup>3</sup> |
| --- | --- | --- | --- |
| <u>Water</u> |  |  |  |
| 7-day diarrhea prevalence | 0.89 (0.70, 1.13) | 1.181 (0.38) | -3.098 (0.97) |
| Length-for-age Z-score | -0.06 (-0.18, 0.05) | 1.000 (0.14) | 0.787 (0.071) |
| Combined EASQ Z-score | 0.15 (0.04, 0.26) | 0.966 (0.0052) | -16.451 (1) |
| <u>Sanitation</u> |  |  |  |
| 7-day diarrhea prevalence | 0.61 (0.46, 0.81) | 0.894 (0.0097) | -144.715 (1) |
| Length-for-age Z-score | -0.02 (-0.14, 0.09) | 0.218 (0.49) | -3.362 (0.99) |
| Combined EASQ Z-score | 0.31 (0.19, 0.43) | 1.002 (<0.001) | 1.255 (0.0028) |
| <u>Handwashing</u> |  |  |  |
| 7-day diarrhea prevalence | 0.60 (0.45, 0.80) | 1.007 (0.058) | -5.434 (0.97) |
| Length-for-age Z-score | -0.07 (-0.18, 0.04) | 0.99 (0.12) | 0.393 (0.27) |
| Combined EASQ Z-score | 0.29 (0.19, 0.39) | 0.996 (<0.001) | 1.068 (0.056) |
| <u>WSH</u> |  |  |  |
| 7-day diarrhea prevalence | 0.69 (0.53, 0.90) | 1.246 (0.21) | -24.749 (1) |
| Length-for-age Z-score | 0.02 (-0.09, 0.19) | 1.155 (0.27) | 0.491 (0.2) |
| Combined EASQ Z-score | 0.24 (0.14, 0.35) | 0.987 (<0.001) | -0.922 (0.85) |
| <u>Nutrition</u> |  |  |  |
| 7-day diarrhea prevalence | 0.64 (0.49, 0.85) | 0.764 (0.0041) | -134.724 (1) |
| Length-for-age Z-score | 0.25 (0.15, 0.36) | 0.993 (<0.001) | -0.208 (0.59) |
| Combined EASQ Z-score | 0.28 (0.18, 0.37) | 1.000 (<0.001) | 0.075 (0.47) |
| <u>Nutrition + WSH</u> |  |  |  |
| 7-day diarrhea prevalence | 0.62 (0.47, 0.81) | 1.037 (0.0043) | -130.676 (1) |
| Length-for-age Z-score | 0.13 (0.02, 0.24) | 1.037 (0.0057) | -3.369 (1) |
| Combined EASQ Z-score | 0.37 (0.27, 0.46) | 1.002 (<0.001) | -0.141 (0.58) |

Note: WSH = Water/Sanitation/Handwashing, EASQ = Extended Ages and Stages Questionnaire
<sup>1</sup>Effect estimates are prevalence ratios with 95% confidence intervals for 7-day diarrhea prevalence and mean differences with 95% confidence intervals for length-for-age Z-score and combined EASQ Z-score.
<sup>2</sup>Coefficient (p-value). P-value <0.05 indicates that the coefficient is significantly different from 1, which suggests that the average prediction from the causal forest is correct.
<sup>3</sup>Coefficient (p-value). P-value <0.05 indicates the presence of heterogeneity from an omnibus test.

### Sorted Group Average Treatment Effects

After predicting individual treatment effects using trained causal forests, ranking individuals based on the magnitude of their individual treatment effects, and dividing the population into terciles, there were differences between tercile average treatment effects only for the EASQ Z-score outcome comparing Sanitation to Control, consistent with the findings from the omnibus test (Figure 2). Comparing Sanitation to Control, children in Tercile 1 (those with the least expected benefit) were predicted to have no treatment effect for EASQ Z-score (0.02 SD, 95% CI: -0.14, 0.19), while children in Tercile 2 and Tercile 3 had significantly larger treatment effects (Tercile 2: 0.39 SD, 95% CI: 0.23, 0.56 and Tercile 3: 0.51 SD, 95% CI: 0.35, 0.67); p-values remained significant after adjusting for multiple testing. All terciles for Handwashing vs. Control on EASQ Z-score had treatment effects statistically different from zero, suggesting all Terciles benefitted from the intervention; although the magnitude of the treatment effect was highest in Tercile 3 (0.41 SD; 95% CI: 0.26, 0.57), it was not significantly different from Tercile 1 (adjusted p-value = 0.14). The monotonic increase in tercile-specific treatment effects for Sanitation vs Control and Handwashing vs. Control for EASQ Z-score outcomes suggests that we were powered to rank some tercile-specific treatment effects (even though there were no differences between terciles with Handwashing vs. Control), while the lack of monotonicity for LAZ comparing Water to Control (Figure 2) suggests we may not be powered to predict tercile-specific treatment effects for this treatment-outcome comparison.

**Figure 2.**
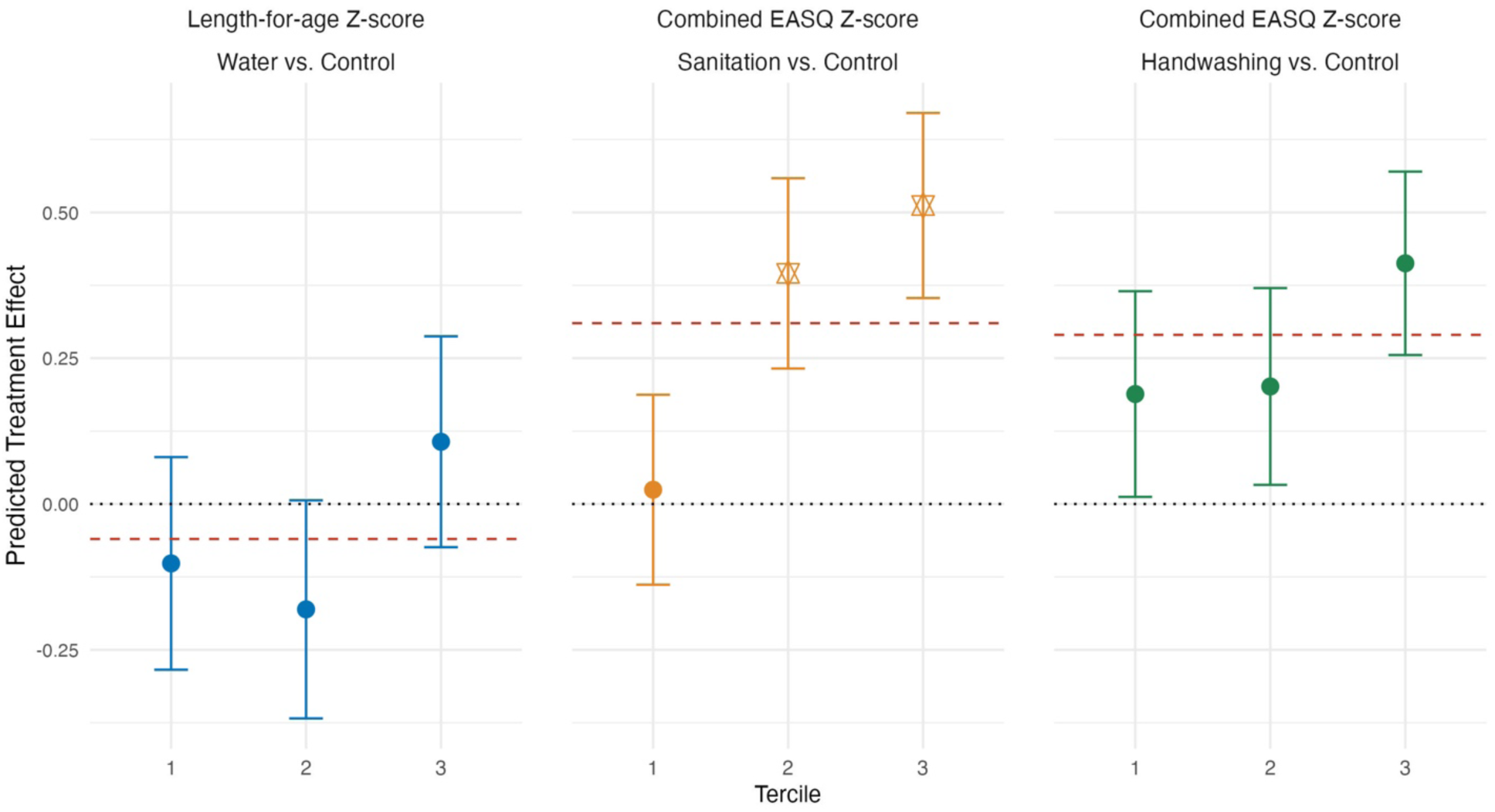
Sorted group average treatment effects (GATES) among treatment-outcome combinations with significant overall heterogeneity. Note: Starred point estimates are significantly different from Tercile 1 at p < 0.05 (adjusted for family-wise error rate using the Romano-Wolf correction). Red dashed lines indicate average treatment effects from the original trial. Length-for-age Z-score and combined EASQ Z-score was measured in index children two years after intervention implementation and 7-day diarrhea prevalence was measured in all children <3 years two years after intervention implementation.

### Variable importance and classification analysis

Comparing the effect of the Sanitation between terciles for the EASQ Z-score outcome, we found differences in baseline characteristics important for predicting heterogeneity (Figure 3). Variables related to animals and animal behaviour in the household, proxies for socioeconomic status, and remoteness were among the most important variables for predicted treatment effect heterogeneity (Figure 3). Households that reported that chickens “always” go inside the house (compared to households reporting that chickens went inside the house “sometimes” or “never”/did not own chickens) was the most important variable, and comparing differences in values between Tercile 1 and 3 (4% vs. 85%) in the CLAN suggests that households where chickens were reported to always go inside the house had higher child development gains from receiving the Sanitation intervention. The number of chickens in the household and compound were important variables; households in Tercile 3 reported more chickens in both, but the differences between terciles were not significant in a parametric test. The second most important variable was whether animal feces were observed in the child’s play area or not (more likely to be observed in Tercile 3; 84% vs. 18%).

**Figure 3.**
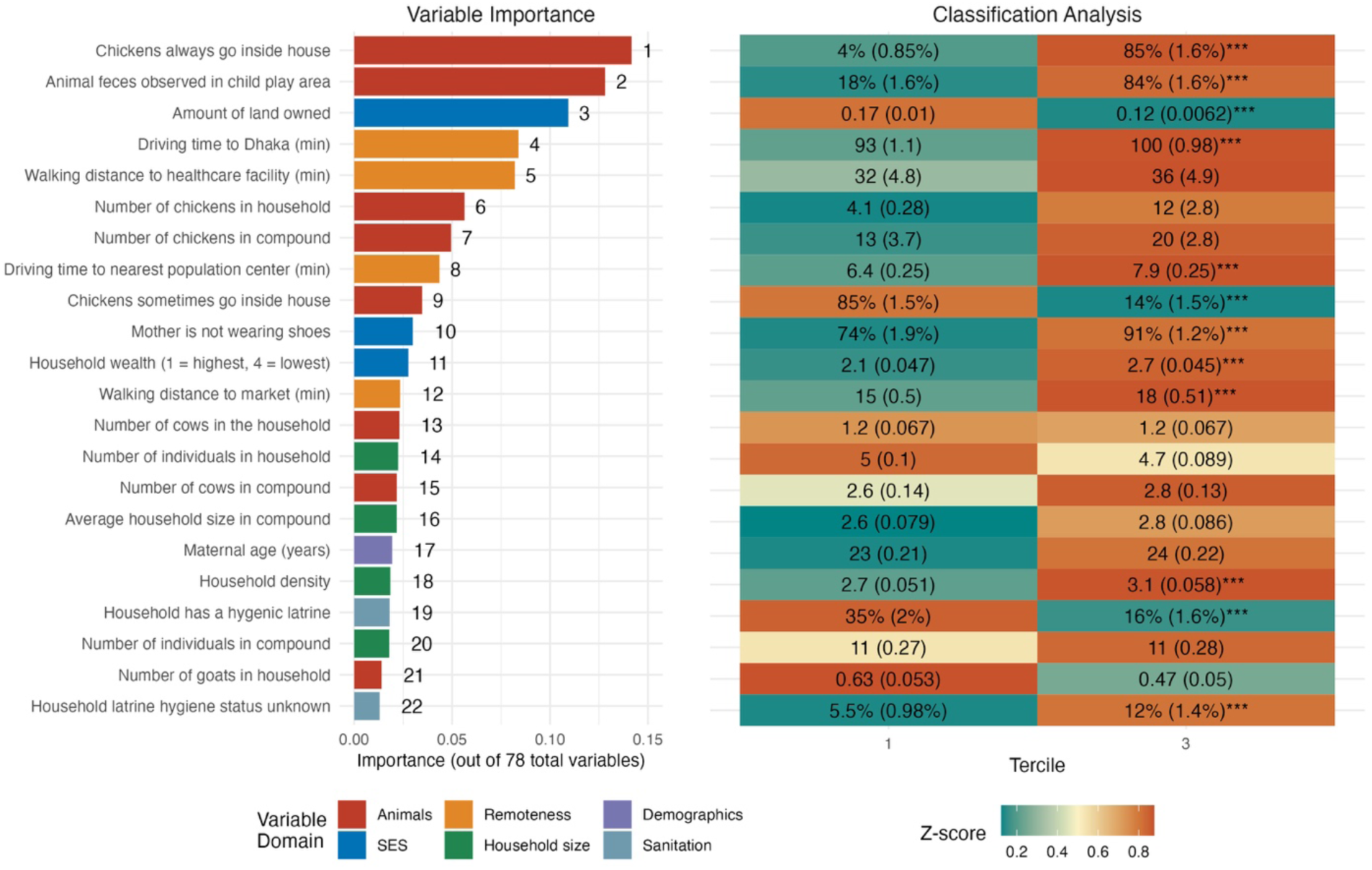
Variable importance and classification analysis for the conditional average treatment effect on combined EASQ Z-score, comparing Sanitation to Control. Note: Variable importance is ordered by a weighted sum of how many times a variable was used for a split in the forest and the depth in the forest the split occurred. For the classification analysis, values are variable means or proportions (standard errors) and colors represent Z-scores within each variable distribution. *** p < 0.001, ** p < 0.05, * p < 0.1 comparing the difference in mean value or difference in proportions, after correction for multiple comparisons using the Bonferroni method.

In terms of household socioeconomic proxies, we found that households that benefited more from the Sanitation intervention were owned less land (#3), less wealthy (#11), and had a higher household density (#18). They were also further from Dhaka (#4), any population center (#8), or a market (#12). Those that benefitted from the Sanitation intervention were also less likely to own a hygienic latrine at baseline (16% owned in Tercile 3 vs. 35% in Tercile 1), although this variable was less important than others (#19).

### Best linear prediction

Using the top five variables in a linear prediction model of *τ* (Figure 4) revealed that variables were similarly directionally associated with treatment effect heterogeneity in the effect of Sanitation on EASQ Z-score, except for driving time to Dhaka. Assuming the linear combination of important variables in predicting heterogeneity to be an accurate specification, we found that chickens always going inside the house was a strong predictor of larger treatment effects on EASQ Z-score in the Sanitation intervention compared to Control, and animal feces observed in the child play area was smaller in magnitude, similar to the results from the variable importance analysis.

**Figure 4.**
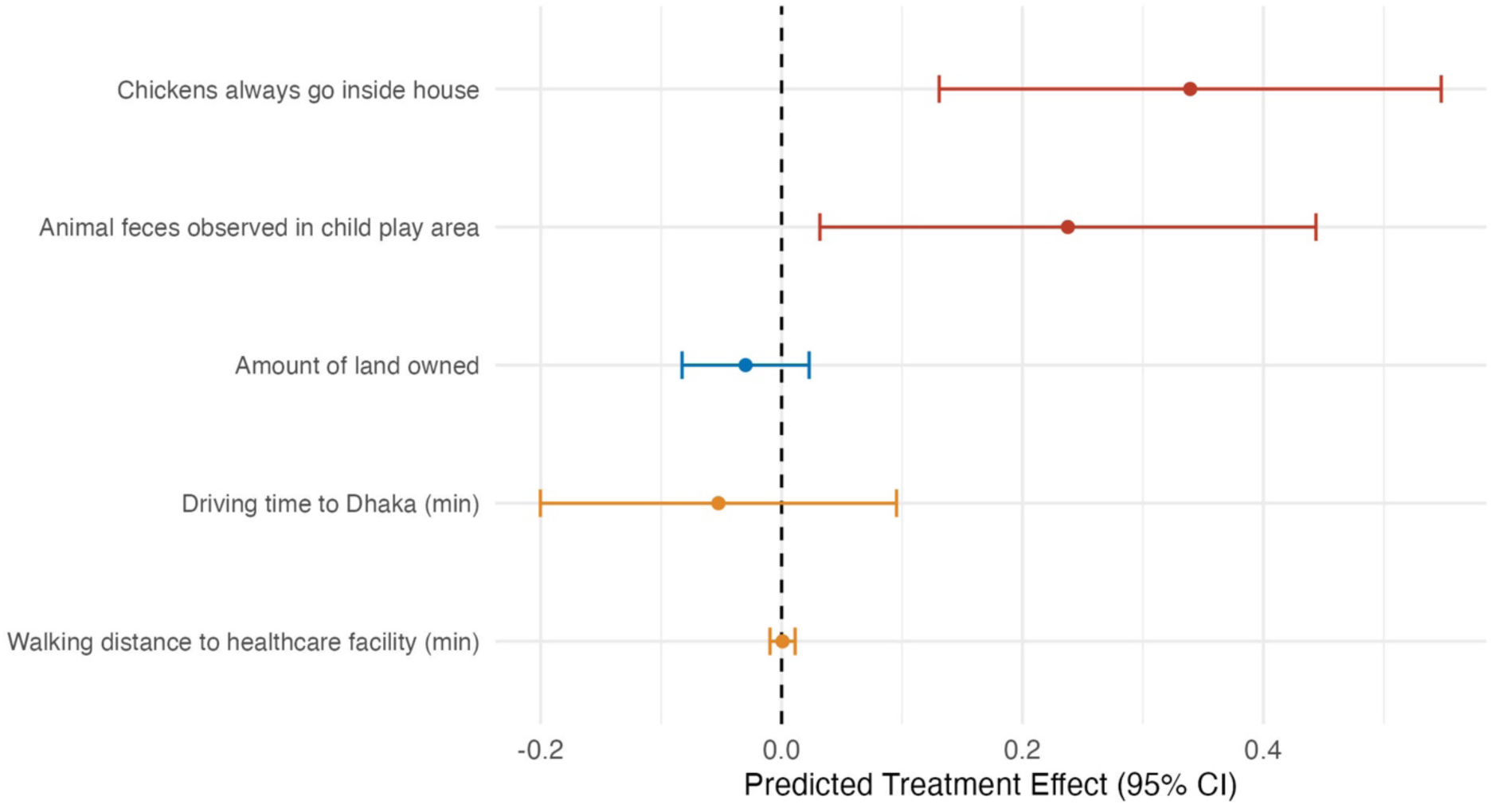
Best linear prediction of the conditional average treatment effect (CATE) for the Sanitation arm compared to Control on combined EASQ Z-score using the top five variables from causal forest. Note: For continuous variables, the point estimate compares the predicted treatment effect for those in the 75th percentile of the variable distribution compared to those in the 25th percentile.

## DISCUSSION

In this study, our goal was to determine if there was detectable treatment effect heterogeneity along easily screenable household characteristics at baseline in a population-based water, sanitation, handwashing, and/or nutrition intervention for different outcomes related to child health using data-adaptive machine learning methods. We did not detect heterogeneity in any treatment arms for the prevalence of 7-day diarrhoea or length-for-age Z-score, showing that children from all backgrounds benefit equally from these interventions in situations where the overall population showed improvements (i.e. the interventions that included nutrition for growth and all interventions for diarrhoea except Water). We found heterogeneity in the effects of receiving sanitation improvements on child development, where poorer households located in more remote areas and potentially with higher levels of animal fecal contamination had the highest expected benefit.

We may not have seen heterogeneity in the impact of the intervention on diarrhoea or growth for several reasons. First, the simple explanation is that there was no heterogeneity for these interventions in this population, as measured by these outcomes, meaning that the treatment effect generated from the original trial is the expected effect for an individual regardless of their covariate values. This suggests that irrespective of household characteristics, children receiving the same intervention benefited equally from that intervention. In the arms containing the nutrition intervention, it is plausible that there was no heterogeneity since the intervention dose was more consistent (while for the WASH interventions the degree to which the beneficiary was exposed due to their use of hardware and related behaviour change was more variable and potentially unknown) and adherence overall was high across arms and easily measured (number of sachets consumed)(12). This is corroborated by a recent analysis using individual participant data from 14 RCTs of small-quantity lipid-based nutrition supplements, including WASH Benefits, where no heterogeneity was found in child growth across all study-level characteristics; the only two individual characteristics that demonstrated effect modification were sex and birth order, neither of which were used in our analysis because at baseline the index children were *in utero*, and we were interested in household characteristics (31). Heterogeneity may also have been obscured by not accounting for the temporality of outcome measurements. Focusing on baseline characteristics as predictors of heterogeneity, our algorithm incorporated season at baseline (time-invariant) to understand if the timing of intervention implementation in regard to rainy versus dry season produces heterogeneous effects, which we did not find to be the case.

However, two recent articles found that interventions in the trial had heterogeneous treatment effects using season of outcome measurement (time-varying), particularly when diarrhoea was measured in the rainy season compared to when diarrhoea was measured in the dry season(17,18). Indeed, diarrhoeal prevalence was not accurately predicted for most of our causal forests. This suggests that season during which outcomes are measured should be considered in future analyses of heterogeneity.

We did find heterogeneity in the effect of the Sanitation intervention on the combined EASQ Z-score, one assessment of child development measured in the main trial. The Sanitation intervention included installing or upgrading all latrines in the compound to hygienic latrines (which have a concrete slab and functional water seal) and providing tools, such as a small hoe for feces removal and child potties, to all households in the compound for the management of animal and child feces around the compound. Some household characteristics related to baseline sanitation were associated with the intervention having greater effects on child development. In particular, these were household characteristics that indicated worse sanitation at baseline such as not owning a hygienic latrine, addressed through hardware provided by the intervention, and if animal feces were observed in the children’s play area, addressed through provided hardware (a feces removal tool) and behaviour change (messaging from community health promoters to remove animal feces from the compound). Thus, one potential pathway by which the intervention may have been able to reduce exposure to fecal pathogens more for certain households and improve cognitive outcomes is by providing tools for the safe removal of animal feces and a latrine for safe disposal. However, environmental sampling at multiple timepoints revealed that fecal contamination measured on child hands or in soil, stored water, or food was no different in the Sanitation arm compared to the Control arm (32). The most important variable associated with intervention effects was if the households reported that chickens always went inside the household and indoor environmental samples were not collected(33), so we are not able to assess if there were heterogeneous effects of the treatment on fecal contamination in the indoor environment. Observational analyses have found that the practice of keeping chickens indoors at night is associated with higher levels of environmental enteric dysfunction and lower growth in Bangladesh (34) and Ethiopia (35). The study in Ethiopia found that chicken ownership was associated with better linear growth, however, keeping chickens inside was associated with a reduction of these benefits. Exposure assessment analyses from children < 3 years in the Control and N + WSH arms determined that contaminated soil was the primary pathway by which children younger than 3 years old were exposed to fecal bacteria (36), but exposure to contamination from indoor floors was not assessed (33).

The Sanitation intervention effects on child development were also predicted to be higher in compounds with smaller land area and higher household density. An observational study using the WASH Benefits Bangladesh cohort found higher latrine coverage within 50 metres was associated with lower fecal contamination in the environment, and with a larger effect in higher population density areas (37). Limited land ownership and high household density may also be proxies for low socioeconomic status, a known risk factor for child development (38). Indeed, one study in Bangladesh found land ownership to be the best indicator of socioeconomic status associated with adverse outcomes from arsenic ingestion (39). We can also corroborate this with process outcomes from the study, where socioeconomic status modified intervention uptake in the Sanitation arm (19); households in the lower wealth quintiles had larger increases in the prevalence of indicators for more hygienic sanitation (e.g. no visible feces on latrine floor, reported use of child potty) from baseline to endline compared to higher wealth quintiles, comparing Sanitation to Control. Socioeconomic status is a strong risk factor for poor child development (38) and less wealthy households may have been more impacted by the Sanitation intervention after accounting for take-up. This was the case for a study in rural Nigeria, which found heterogeneous impacts of a community-led total sanitation intervention across community wealth, with less wealthy communities having higher treatment effects, over-and-above intervention take-up (40).

Another possibility is that child development was heterogeneously affected by pathways other than those related to sanitation. One hypothesis for why the WASH interventions had strong effects on child development but not on child growth is related to reductions in maternal depression and stress caused by the intervention, which likely increased responsive caregiving (41). The Sanitation intervention was the only intervention that was delivered to every household in the compound where the target child lived, so the general cleanliness of the mother’s environment may have improved, improving maternal mental health and thus child development heterogeneously depending on the baseline characteristics of the compounds. Given that sanitation insecurity can affect multiple psychosocial dimensions related to maternal well-being (42,43), fecal contamination and reduction of diarrhoea could be considered only partial indicators of WASH intervention success (44). It may be important to also look at how the provision of basic sanitation services can improve outcomes beyond fecal contamination and clinical diarrhoea among those most vulnerable to poor child development. Similar arguments could be made for the treatment of water and supplies for handwashing, but few households owned water treatment or a handwashing station at the beginning of the intervention and thus we would be unable to detect any heterogeneity for these dimensions.

We also found heterogeneity for EASQ Z-score only in the individual Sanitation intervention and not the combined WSH or Nutrition + WSH intervention. One explanation could be that the combined interventions were able to address contamination pathways more completely. That is, if we assume that the Sanitation intervention addressed pathways affected by sanitation and that the Handwashing intervention addressed pathways affected by handwashing, addressing multiple pathways at once may have produced more homogenous effects, whereas addressing only certain pathways produced heterogeneous effects. Infectious diseases modeling work using WASH Benefits data has concluded that addressing multiple exposure pathways at once is the most important factor in the reduction of clinical diarrhoea (21). However, given the absence of additive effects in the main trial both for environmental data (33) and clinical outcomes (12), and that the Sanitation intervention could have more completely addressed contamination pathways than the individual water and hygiene arms, this may not explain the differences we found in this analysis.

There were some limitations to our analysis. We focused on variables related to household conditions assessed or observed at baseline, which occurred when the child was *in utero*, to make inferences about households to target for these specific interventions. The selected variables were primarily related to household WASH characteristics; a set of variables related to biological factors or nutritional practises may have led to more observed heterogeneity in the nutrition intervention. Also, although the WASH-Benefits study had a rich baseline questionnaire, it may be that the values of these variables changed after baseline and did not represent a child’s exposure to the selected potential treatment effect modifiers over the first two years. The alternate explanation is that there was heterogeneity along the chosen baseline variables, but it was too small to detect with the sample size available for analysis. This may be especially true for the outcomes where the average treatment effects were small or null (i.e. diarrhoea prevalence and LAZ). We did not hypothesize the treatment to be harmful for any subgroups, and while there may have been small subgroups who benefitted, we may have been underpowered to detect this. However, it is possible that any heterogeneity that could be detected with a larger sample size would not be meaningful enough to influence targeting for WASH interventions. Another limitation of our analysis was that we decided posthoc to reduce the number of predicted groups from quintiles to terciles to improve power. This may have reduced differences between groups, however we still found differences in baseline characteristics between groups among causal forests with significant heterogeneity. If these methods are of interest to future researchers, a larger sample size would be advisable, which may be easier when using administrative data or with interventions already planned to cover a large area, in addition to collecting rich baseline data.

## CONCLUSION

Baseline characteristics of households did not predict heterogeneity for the presence of diarrhoea or growth at two years, suggesting that children from all backgrounds can benefit from these interventions, when an overall effect is present. Children born into poorer and more remote households with worse fecal contamination from animals and chickens always allowed in the house were predicted to gain higher development scores from the sanitation intervention alone. Identifying such subgroups may allow for targeting in resource-constrained settings; given that universal WASH is a human right and requires universal coverage for maximal effects, targeting may rather take the form of prioritizing households during universal rollout or targeting promotion or maintenance checks. Prioritizing households with these characteristics for sanitation interventions may result in higher impacts, especially in situations of budgetary constraints and ahead of more transformative, community-based solutions. The use of causal forests and other machine learning algorithms allows for the exploration of subgroups defined by complex combinations of covariates not possible in prespecified analyses, which can be a useful, data-adaptive way to uncover drivers of treatment effect heterogeneity. Given that trials are resource-intensive, utilizing these tools from the fast-moving data science field and leveraging randomization can assist with maximally understanding who the intervention did and did not help, or if no heterogeneity exists overall, for future intervention design and delivery in resource-constrained settings.

## Data Availability

Outcome data and some of the household characteristics are publicly available on OSF. The remaining data are available upon reasonable request (contact Andrew Mertens).

https://osf.io/tprw2/

## Acknowledgements

Thank you to Ben Arnold and Francois Rerolle for providing scripts to extract travel time data. Thank you to Anna Nguyen and Jessica Grembi for providing data extracted from remote sensing for consideration in the predictor dataset. Our extreme gratitude to all those involved in the WASH Benefits studies from conception to completion.

## Ethics approval

The WASH-Benefits Bangladesh trial was approved by Ethical Review Committee at The International Centre for Diarrhoeal Disease Research, Bangladesh (PR-11063), the Committee for the Protection of Human Subjects at the University of California, Berkeley (2011-09-3652), and the institutional review board at Stanford University (25863).

## Author contributions

CH, LHK, and ANM conceived the study. CH conducted the analysis, interpreted the results, and drafted the original manuscript. ANM contributed to the data analysis, results interpretation, and original manuscript revisions. ANM, LHK, LCHF, AE, JMC, FT, MR, SP, and SPL reviewed the manuscript and provided substantive feedback and revisions. All authors approved the final version of the manuscript and agree to be accountable for all aspects of the work. CH is the guarantor of the study.

---

**Supplementary Table 1.**
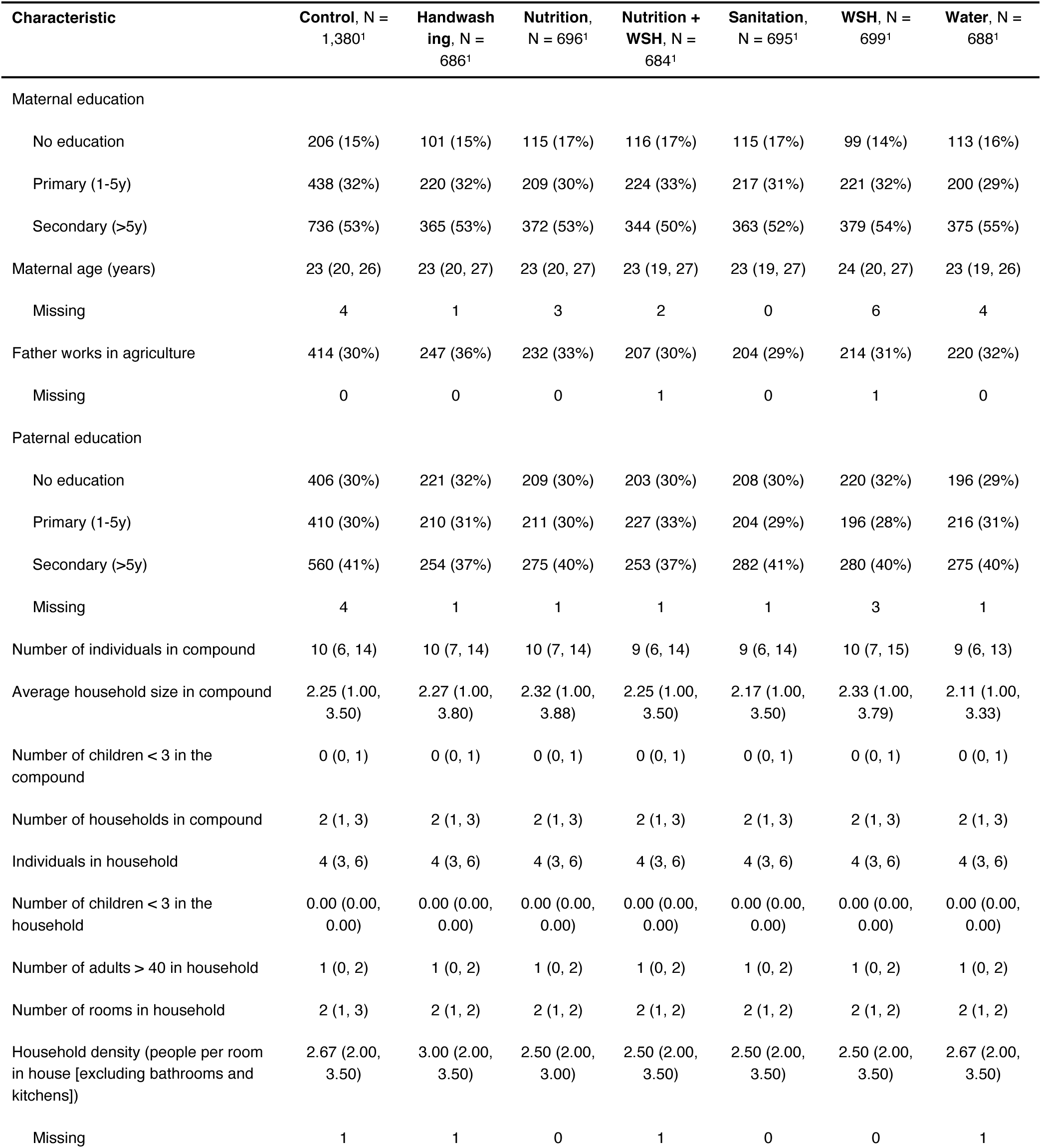
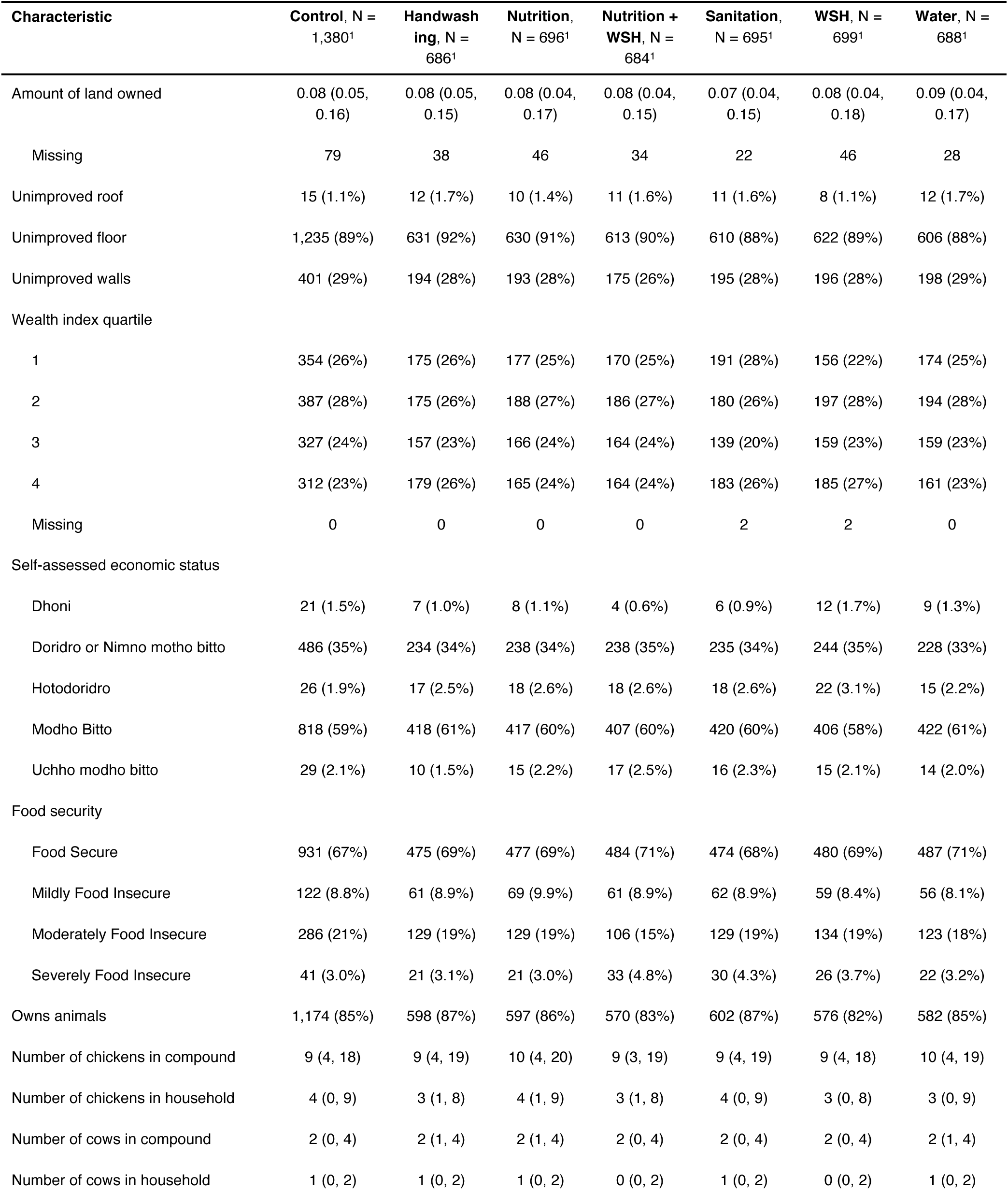
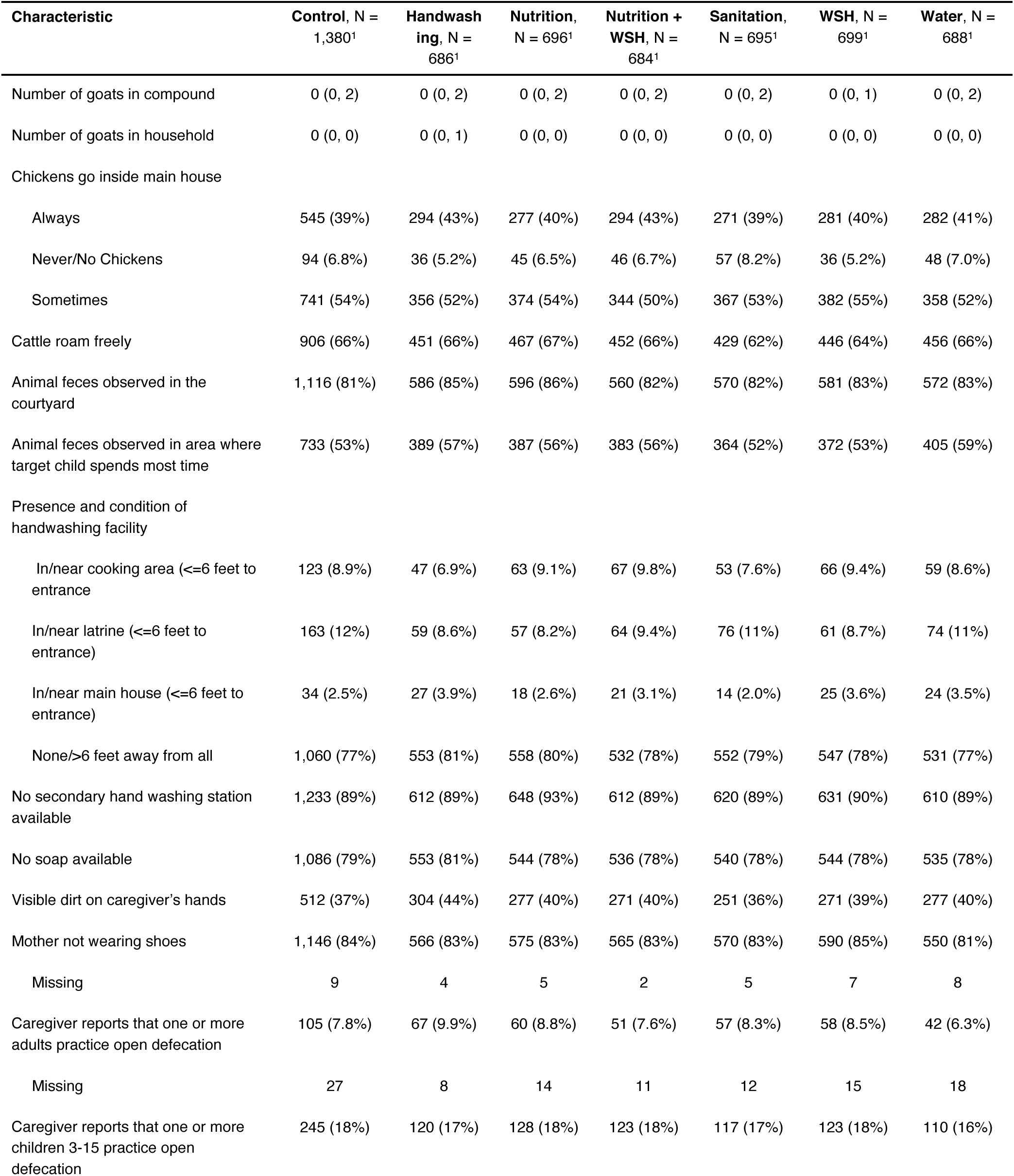
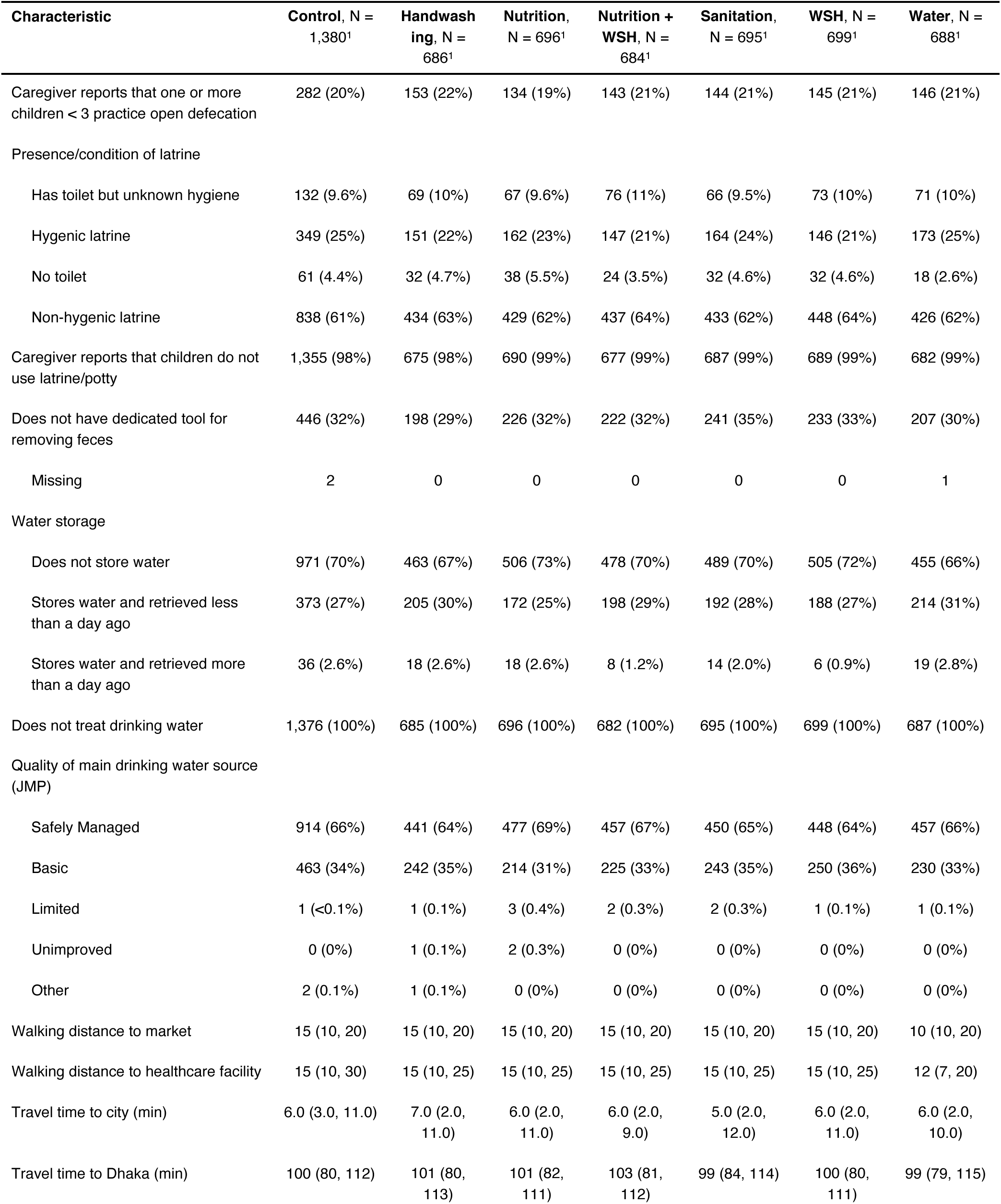
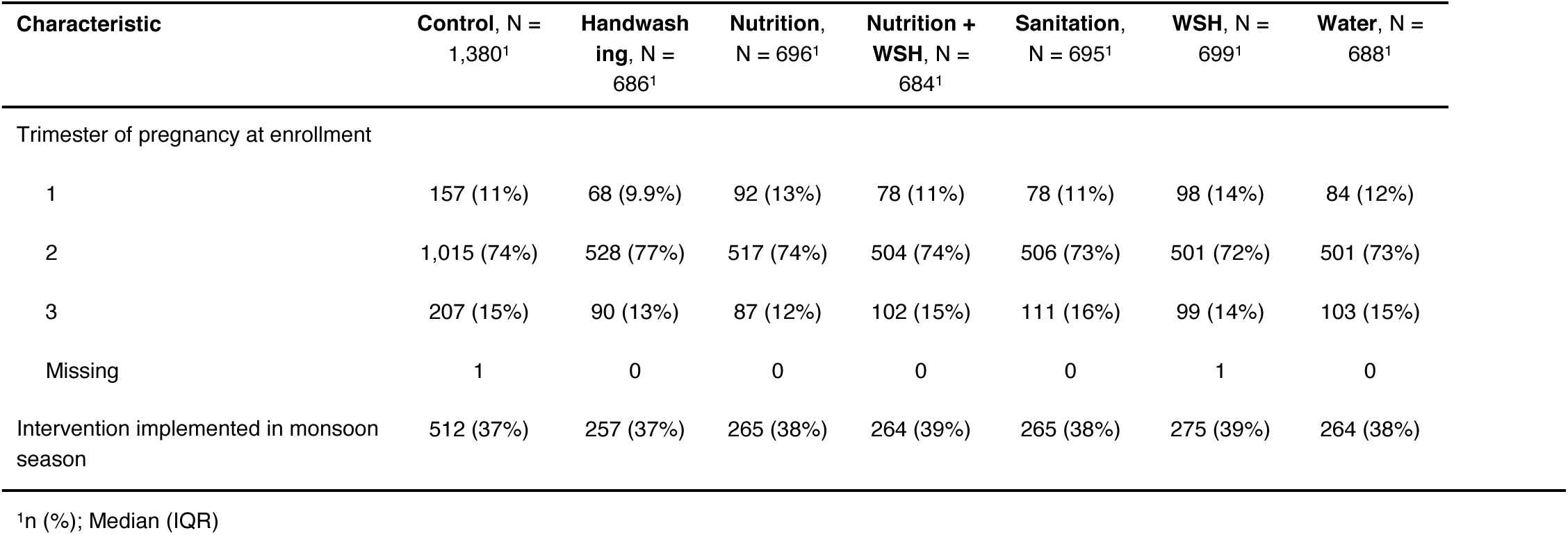
Characteristics of households at baseline, by treatment arm.

**Supplementary Figure 1.**
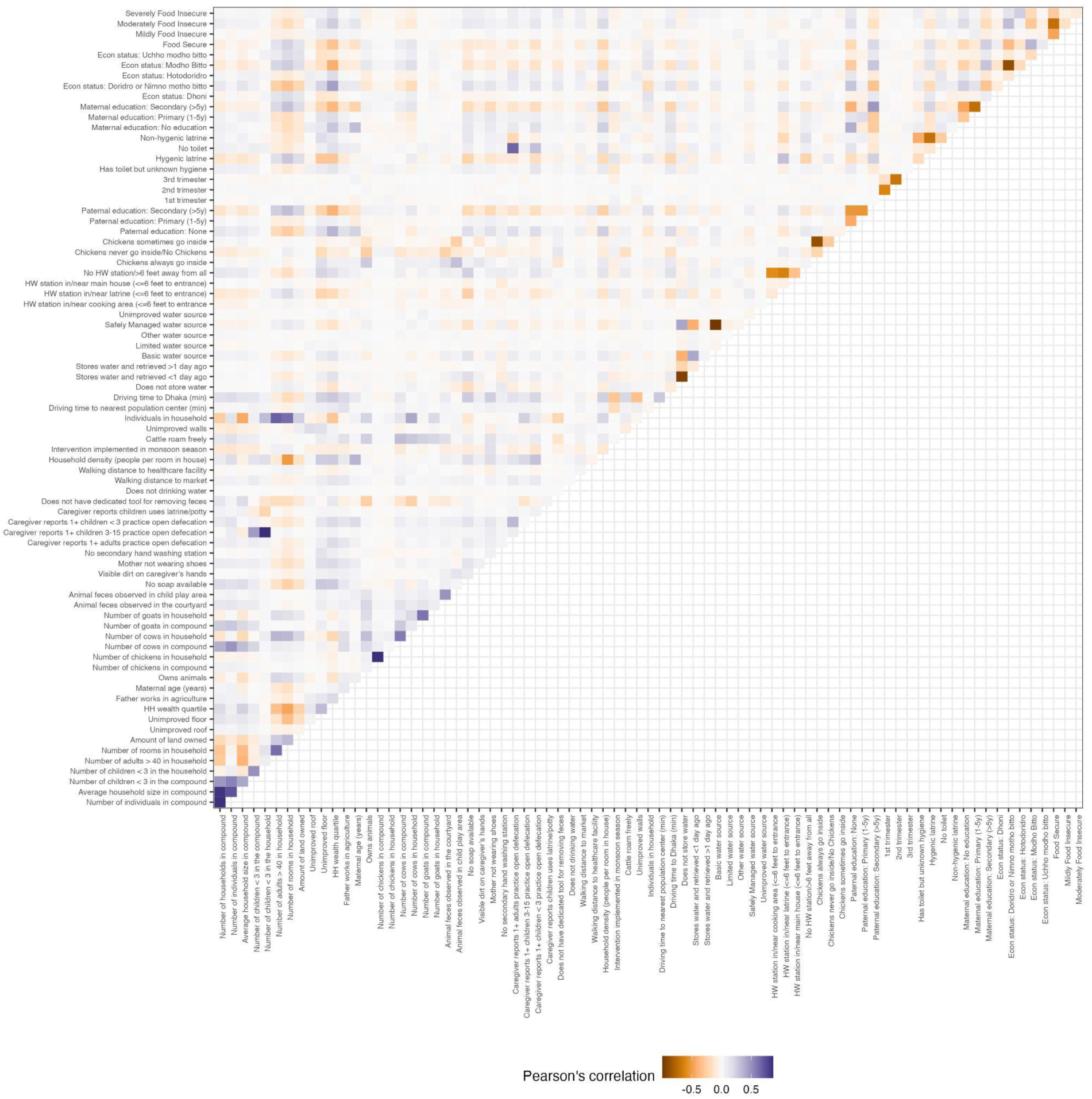
Plot of pairwise correlations for baseline variables

